# COVID-19 in Children with Brain-Based Developmental Disabilities: A Rapid Review

**DOI:** 10.1101/2020.05.01.20088021

**Authors:** Michèle Dugas, Valérie Carnovale, Andrée-Anne Poirier, Benoit Mailhot, Becky Skidmore, Lena Faust, Carrie Costello, Donna Thomson, Annette Majnemer, Dan Goldowitz, Steven P. Miller, Annie LeBlanc

**Author notes:** **Corresponding author:** Dr. Annie LeBlanc PhD. VITAM, Centre de recherche en santé durable, 2601, chemin de la Canardière (bureau G-2639), Québec (Québec) CANADA, G1J 2G3.

## Abstract

**Background:** The prevalence of symptomatic COVID-19 in children remains low to date. In just a few months, COVID-19 has affected millions of people worldwide, and as of the date of this publication, the pandemic continues. Based on the current available evidence, children do not appear to be at higher risk of contracting COVID-19 than adults. However, children with neurological and neuromuscular conditions are vulnerable to the respiratory complications of other viral infections.

**Objectives:** To assess whether children with brain-based developmental disabilities were more likely to develop COVID-19 and have complications or poorer outcomes following infection.

**Methods:** We conducted a two-week rapid review on studies with primary data regarding children aged between zero and 18 years old with brain-based developmental disabilities, or who were at risk of developing such disabilities, with confirmed or suspected COVID-19. We performed our literature searches on April 18, 2020.

**Results:** Our search strategy identified 538 individual records, of which four were included in our review. Of the 50 COVID-19 pediatric patients reported in the included studies, a total of seven children were at risk of developing brain-based disabilities. Symptoms ranged in severity. However, generally, patients were discharged or saw improvements in their symptoms by the end of the study period. No deaths were reported.

**Discussion:** Our study highlights a knowledge gap regarding the impact of COVID-19 in children with brain-based developmental disabilities.

## Background

According to the World Health Organization, SARS-CoV-2 has infected close to three million and caused the death of over 200 000 individuals worldwide, as of April 26th, 2020 (1). The prevalence of COVID-19 in children remains low to date. Of the 23,082 cases reported in Canada (April 23rd, 2020), 1,055 (4.6%) were in patients aged 19 years old and under (2). Of those, 14 were hospitalized, and two were admitted to Intensive Care Unit, with no deaths being reported. Data gathered for the U.S. between February 12th, 2020 and April 2nd, 2020 indicated that of the 149,760 cases, 1.5% (2,572) were children, of which 13% had underlying conditions, 5.7% were hospitalized, and included three deaths (3). As of February 11th 2020, China had reported that of 44,672 confirmed cases in mainland China, 965 (2.1%) were children, with one reported death (4). As of April 24rd, 2020, a total of 150,383 cases were reported in Germany, of which 2,476 (2%) were under 10 years old, and 6,209 (4%) were aged between 10 to 19 years old (5). Numbers of hospitalizations and deaths were not available. Finally, as of March 12th, 2020, 7,755 cases were reported in the Republic of Korea, of which 480 (6.2%) were pediatric, with no deaths being reported (6).

Based on the current available evidence, children do not appear to be at higher risk of contracting COVID-19 than adults (7). However, children with comorbidities may be vulnerable to severe COVID-19 disease (8). Indeed, children with neurological and neuromuscular conditions had an increased risk of respiratory failure when hospitalized with influenza, another viral infection (9). Symptom manifestation in children appears to be milder than in adults (10, 11). Further, the incidence in individuals under 19 years of age has been quite low (8, 10, 12) and children could even be less susceptible to the COVID-19 disease (7). To the best of our knowledge, no review has searched for the impact of COVID-19 on children with brain-based disabilities affected by COVID-19.

## Objectives

This rapid review was commissioned by the Strategy for Patient-Oriented Research (SPOR) funded CHILD-BRIGHT Network, an innovative pan-Canadian network that aims to improve life outcomes for children with brain-based developmental disabilities and their families (https://www.child-bright.ca/). Concerned with the potential impact of the novel coronavirus on children with brain-based developmental disabilities, they requested support from the SPOR Evidence Alliance to conduct a rapid review within a two-week period, on the topic. Thus, this review aimed to answer the following questions:

1. Are children with brain-based developmental disabilities more likely to develop COVID-19?
2. Are children with brain-based developmental disabilities more likely to develop complications due to COVID-19?
3. Are children with brain-based developmental disabilities more likely to have a poorer prognosis once they develop COVID-19?

We engaged with a panel of knowledge users (patients, caregivers, clinicians, decision makers) and researchers from the CHILD-BRIGHT Network throughout the review process, from question development, literature search, interpretation and writing of results, and dissemination of findings.

## Methods

We conducted the rapid review based on the proposed methodology guide of the Cochrane Rapid Reviews Methods Group (13). We report our results based on the Preferred Reporting Items for Systematic Reviews and Meta-Analyses (PRISMA) Statement (14).

### Literature Search

An experienced medical information specialist developed the search strategies through an iterative process in consultation with the review team and the panel of knowledge users. The MEDLINE strategy was peer reviewed by another senior information specialist prior to execution using the PRESS Checklist (15). Using the OVID platform, we searched Ovid MEDLINE^®^, including Epub Ahead of Print and In-Process & Other Non-Indexed Citations, Embase Classic+Embase, PsycINFO, Cochrane Database of Systematic Reviews and the Cochrane Central Register of Controlled Trials. We also searched CINAHL (Ebsco) and Web of Science. All searches were performed on April 18^th^, 2020.

We used a combination of controlled vocabulary and keywords (e.g., “Coronavirus Infections”, “Coronavirus”, “Child”) for the strategies and adjusted vocabulary and syntax across databases. We initially also included vocabulary and keywords specific to brain-based developmental disabilities but removed them after piloting of that strategy yielded no citations to be included in the review. There were no language restrictions on any of the searches but when possible, animal-only records were removed from the results. We limited results to publication years 2019 to the present. Specific details regarding the strategies appear in Appendix 1.

From the included studies and the reviews identified from our searches, we reviewed reference lists for original studies and cross-referenced them with a list of articles provided by content experts from our knowledge users panel. Considering the fast pace at which information becomes available in the context of COVID-19, we developed a grey literature search strategy in consultation with our experienced medical information specialist, which consisted of preprint articles from SSRN and medRxiv (last consulted April 23^rd^, 2020), ongoing trials from the WHO International Clinical Trials Registry Platform (last consulted April 23^rd^, 2020), ongoing reviews from PROSPERO (last consulted April 23^rd^, 2020), and Government or Health organizations’ websites and reports (consulted between April 17^th^ to April 25^th^, 2020) (Appendix 2).

### Eligibility Criteria

We followed the PECO Framework in establishing eligibility criteria (16, 17) (Table 1). We considered any study with primary data that included children aged between zero and 18 with a brain-based developmental disability or at risk of developing such disability with confirmed or suspected COVID-19 (see Appendix 3 for full list).

**Table 1.** PECO Inclusion Criteria

|  |  |
| --- | --- |
| **Population (P)** | Children (18 years and under) with brain-based developmental disabilities (e.g., cerebral palsy, autism, developmental delay, attention deficit hyperactivity disorder, severe impairments) or at risk of developing brain-based disabilities (e.g., premature, congenital heart disease/defect). |
| **Exposure (E)** | COVID-19 |
| **Comparator (C)** | Children with COVID-19 or no comparator |
| **Outcomes (O)** | Any outcome |

### Study Selection and Extraction

Four reviewers individually performed screening for titles, abstracts and then full text using pilot-tested standardized forms (25 and five citations respectively for each level of screening). We developed a standardized extraction form that included study (e.g., authors, country, study design) and case characteristics (e.g., type of disability, age), complications, and any outcome reported. Single reviewers extracted data which was then confirmed by a senior reviewer. We resolved discrepancies through discussion.

### Risk of Bias Appraisal

No risk of bias was performed due to the short turnaround timeline.

### Synthesis

We report data using a narrative approach which includes tables of study characteristics and detailed reporting of case characteristics, complications, and outcomes. Our data synthesis focused on providing a descriptive summary to inform knowledge users.

## Results

### Literature Search

Our search strategy identified 538 individual records. Following the screening of titles and abstracts, we excluded 331 records. After full-text screening of 207 records, we excluded an additional 203 records resulting in a total of four records included in our review (18–21) (Figure 1).

**Figure 1.**
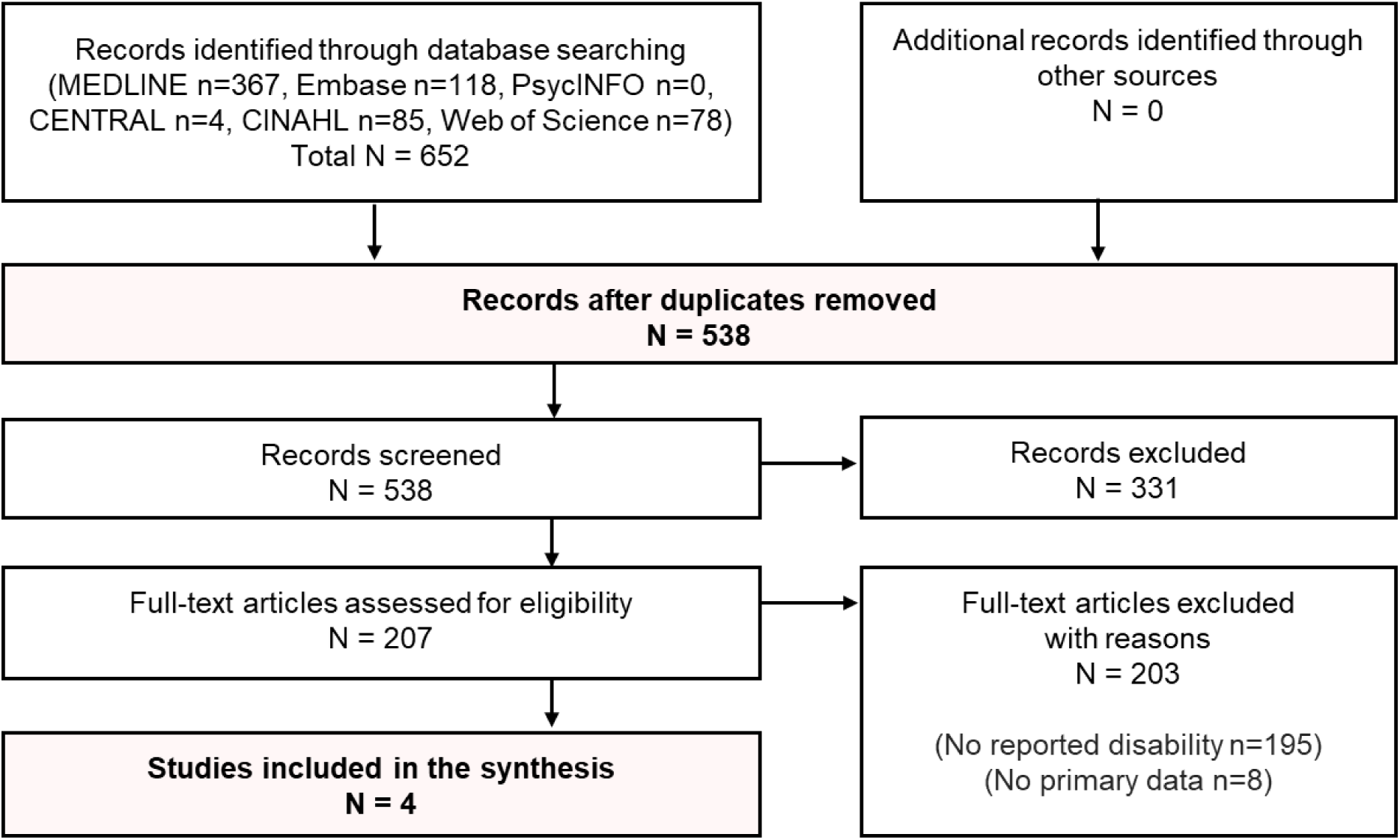
Study Flow Diagram

Our grey literature search did not identify any ongoing trials or preprint articles of research regarding children with brain-based developmental disabilities.

### Characteristics of Included Studies

Of the four included studies, one was a case report (18), one was a cohort study (19), one was a retrospective cross-sectional analysis (20), and one was a retrospective study of medical records (21) (Table 2). Three studies took place in Wuhan, Hubei province, China (19–21) and one in the U.S. (18). The four included studies reported on a total of 80 pediatric patients with 50 infected or suspected of being infected.

**Table 2.** Description of included studies

| Author (year) | Country | Title | Study aims | Study design | Population |
| --- | --- | --- | --- | --- | --- |
| Paret (2020) | USA | SARS-CoV-2 infection (COVID-19) in febrile infants without respiratory distress | Reporting of two cases of COVID-19 in infants | Case report | Infants (n=2) |
| Zeng (2020) | China (Wuhan) | Neonatal early-onset infection with SARS-CoV-2 in 33 neonates born to mothers with COVID-19 in Wuhan, China | Examining neonatal early-onset infection with COVID-19 in 33 neonates born to mothers with COVID-19 | Cohort study | Infants (n=33)<br>Only 3 infected |
| Zheng (2020) | China (Wuhan) | Clinical characteristics of children with coronavirus disease 2019 in Hubei, China | Identifying clinical characteristics of children with COVID-19 | Retrospective cross-sectional study | Children (n=25) |
| Xia (2020) | China (Wuhan) | Clinical and computed tomography features in pediatric patients with COVID-19 infection: Different points from adults | Discussing characteristics of clinical, laboratory, and chest computed tomography in pediatric patients from adults with COVID-19 | Retrospective study of medical records | Children (n=20) |

### Characteristics of Cases

We did not identify any study reporting on children with brain-based developmental disabilities, all included studies reported on children at risk of developing a brain-based developmental disability. Two of the studies included preterm infants (18, 19) while the others included children with congenital heart disease or epilepsy (20, 21).

Detailed information was only available for three of the four studies (18–20) (Table 3). Symptoms for preterm infants ranged from a single fever to overall mild symptoms (18, 19), with one infant developing complications (fetal distress, neonatal respiratory distress syndrome, pneumonia, and suspected sepsis) and requiring resuscitation at birth (19). The preterm infant without complications was discharged in stable condition (18) whereas the infant developing complications saw its condition resolve on day 14 of life (19). Children with congenital heart disease expressed symptoms including cough, dyspnea, fever, and diarrhea and required pediatric intensive care with invasive mechanical ventilation (20). Their symptoms were later partially or significantly alleviated (20).

**Table 3.**
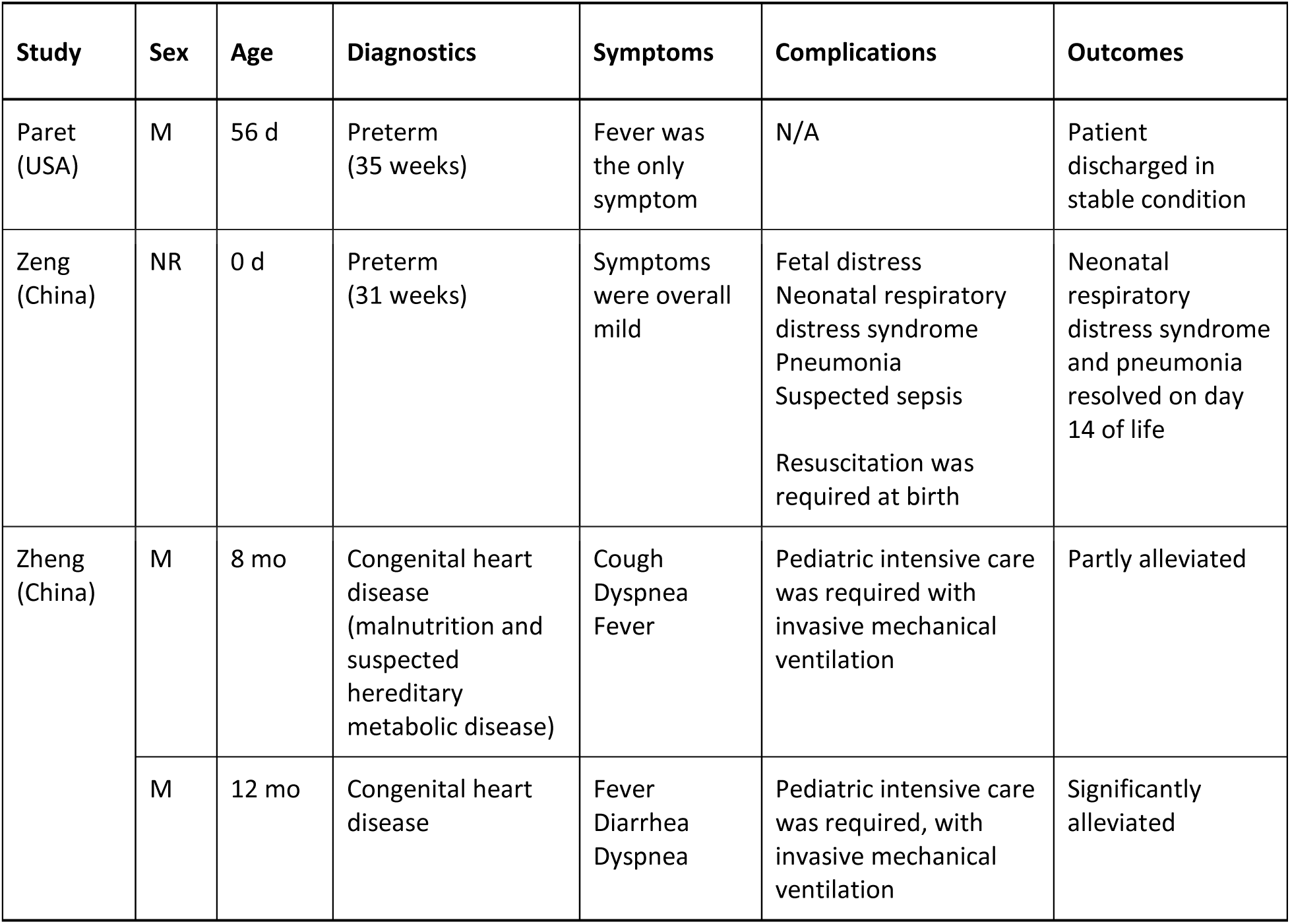
Summary of cases

The fourth study identified one patient with epilepsy (resulting from sequela of previous encephalitis) and two with a history of atrial septal defect surgery but no individual information was available (21). Overall, symptoms were mild, and eighteen of the 20 children were cured and discharged with an average stay of 12.9 days (8-20 days).

## Discussion

The prevalence of COVID-19 in children has been reported as being low. However, we were unable to find information regarding the prevalence of this coronavirus in children with brain-based developmental disabilities. We did not identify any study investigating the effects of COVID-19 in children with brain-based disabilities but did identify four studies discussing the coronavirus disease in children considered at risk for disability (preterm infants or children with some pre-existing medical conditions). Although some of the children in the included studies did develop complications, symptoms were reported as being partially or significantly alleviated or resolved, with most patients being discharged from the hospital and no deaths being reported. These results are in accordance with other studies on COVID-19 in children which reported that symptoms in the pediatric population appeared milder than in adults with the occurrence of death being very rare (11, 12, 22).

Our review highlights the lack of evidence regarding the effects of COVID-19 in children with brain-based disabilities and those with at-risk conditions. Indeed, it appears as though this specific population has been overlooked. The lack of data could be due to the low occurrence rates as well as the milder symptoms which have previously been reported in children (11, 12, 22). In addition, it is possible that due to their milder or absent symptoms, these cases simply do not get reported. However, clinical guidelines from the Centers for Disease Control and Prevention (CDC) clearly state that individuals of any age with underlying conditions, including congenital heart disease, may be at higher risk of developing moderate to severe symptoms from COVID-19 (23). Moreover, UNICEF warns that children with underlying disabilities may be at greater risk of developing complications (24). More data, including the presence of brain-based disabilities and other at-risk conditions in children, are required to have a better understanding of the clinical impacts of COVID-19 on these potentially more vulnerable populations. With that goal in mind, one ongoing review is currently examining comorbidities in the adult and pediatric populations (25).

### Limitations

Our rapid review has limitations. Due to our study design, screenings and data extractions were performed by single reviewers. However, we did perform pilot-testing for each review form to optimize consistency between reviewers. Further, despite our best efforts to identify all relevant studies or relevant documentation, it is possible that some were missed due to the rapid flow at which new information becomes available.

## Implications for Practice and Policy

Our rapid review has identified knowledge gaps in the literature regarding the effects of COVID-19 in children with developmental brain-based disabilities, and those at risk of developing such disabilities. Without data regarding children and high-risk populations, it is difficult for decision-makers to determine the best course of action not only for medical treatment, but also the reintegration of children with disabilities to school and the community with regards to the eventually alleviated confinement measures.

## Data Availability

Data is available upon request to the corresponding author.

## Acknowledgements

The authors would like to thank the panel of knowledge users for their support throughout this review.

